# The epidemiological characteristics of 2019 novel coronavirus diseases (COVID-19) in Jingmen, Hubei, China

**DOI:** 10.1101/2020.03.07.20031393

**Authors:** Qijun Gao, Yingfu Hu, Zhiguo Dai, Feng Xiao, Jing Wang, Jing Wu

## Abstract

**Background:** There is currently a global outbreak of coronavirus disease 2019 (COVID-19),and its epidemic characteristics in the areas where the outbreak has been successfully controlled are rarely reported.

**Objective:** Describe the epidemic characteristics of COVID-19 in Jingmen,Hubei,introduce the local prevention and control experience,and observe the impact of various prevention and control measures on the number of new cases.

**Methods:** All the COVID-19 patients diagnosed in the municipal districts of Jingmen from January 12 to February 29,2020 were enrolled in this study. We described epidemiological data and observed the impact of control measures on the epidemic.

**Findings:** Of the 219 cases (110 men and 109 women), 88 (40%) had exposure to Wuhan. The median age was 48 years (range,2-88 years;IQR,35-60). Thirty-three severe patients with a median age of 66 years(range,33-82 years,IQR,57-76) were treated in intensive care units; out of these patients, 66.7 %(22) were men and 19 (57.5%) had chronic diseases, including hypertension, diabetes, heart failure, stroke, and renal insufficiency. Under the control measures, the number of new patients gradually decreased and nearly disappeared after 18 days. Wearing masks in all kinds of situations prevents most infections and is one of the most effective prevention and control measures.

**Interpretation:** In conclusion,all people are susceptible to COVID-19, and older males and those with comorbid conditions are more likely to become severe cases. Even though COVID-19 is highly contagious,control measures have proven to be very effective, particularly wearing masks,which could prevent most infections.

**Funding:** Supported by the Major Program of Technological Innovation of Hubei Province.

## Introduction

Wuhan, the capital of Hubei province in China, reported an outbreak of atypical pneumonia caused by the severe acute respiratory syndrome coronavirus 2 (SARS-CoV-2) on December 31, 2019, which was then named the coronavirus disease 2019 (COVID-19) by the WHO.^1-4^ Cases spread to other Chinese cities, as well as other countries and regions. The number of cases in Wuhan and other cities in China increased rapidly.^5-7^ On February 29, authorities reported more than 66,000 confirmed cases and 2,727 deaths in Hubei, with most occurring in Wuhan. The outbreak was controlled through the implementation of control policies after January 23. However, the situation has deteriorated outside of China; cases have surfaced in more than 100 countries,^8^ with outbreaks in many nations.^9^

Some articles have reported the epidemiological and clinical characteristics of COVID-19 in Wuhan,^10-13^ but the epidemic characteristics in other cities have rarely been reported. Clinical manifestations do not vary widely, but the epidemiology may differ from region to region. Wuhan once seemed completely out of control: thousands of people were waiting to see doctors, and many people died while waiting for hospital beds. More than 30,000 medical workers nationwide were sent to Wuhan relieve the shortage of medical resources. The situation in Jiangmen was quite different, and strict control measures were put in place when the outbreak began, a successful example of controlling COVID-19. We analyzed the epidemiology of 219 confirmed COVID-19 patients admitted to the First People’s Hospital of Jingmen, which admitted all the confirmed patients within the municipal districts (not including county or county-level cities) of Jingmen, a city in Hubei located about 250 miles from Wuhan.

## Methods

### Patients

For this retrospective study, we recruited patients in the First People’s Hospital of Jingmen from January 12 to February 29, 2020. All the COVID-19 patients diagnosed in the municipal districts of Jingmen were enrolled in this study. The government closed all city entrances on January 26, but before that, some patients arrived from other locations. The study was approved by the First People’s Hospital of Jingmen Ethics Committee. The patients’ written informed consent was obtained before data was collected.

### Procedures

Epidemiological data were collected from patients’ medical records. If more information was needed, we communicated with attending health care providers, patients, and their families. Throat-swab specimens from the upper respiratory tract were obtained for real-time reverse transcription polymerase chain reaction (RT-PCR). The Wuhan Institute of Virology provided the RT-PCR detection reagents.

### Outcomes

We described epidemiological data and observed the impact of control measures on the epidemic.

#### Statistical analysis

Statistical analysis was performed with SPSS 19.0. Continuous variables were described using mean, median,interquartile range (IQR) values and categorical variables were described as frequency rates and percentages.

## Results

### The demographic characteristics

The 219 patients were aged 2–88 years old with 50.2% of the sample being men. The median age of the patients was 48 years (IQR, 35-60,figure 1). Including 33 patients in the intensive care unit (ICU), the median age of the patients was 66 years(IQR, 57-76), ranging from 33–82 years old, with 22 (66.7%) being men and 19 (57.5%) patients having chronic diseases, including hypertension, diabetes, heart failure, stroke, and renal insufficiency. By the end of January 29, eleven patients had died (Six patients had uremia with long-term dialysis), and six of them were men. The average age of the patients who died was 58.3, with a range from 35–78 years old.

**Figure 1:**
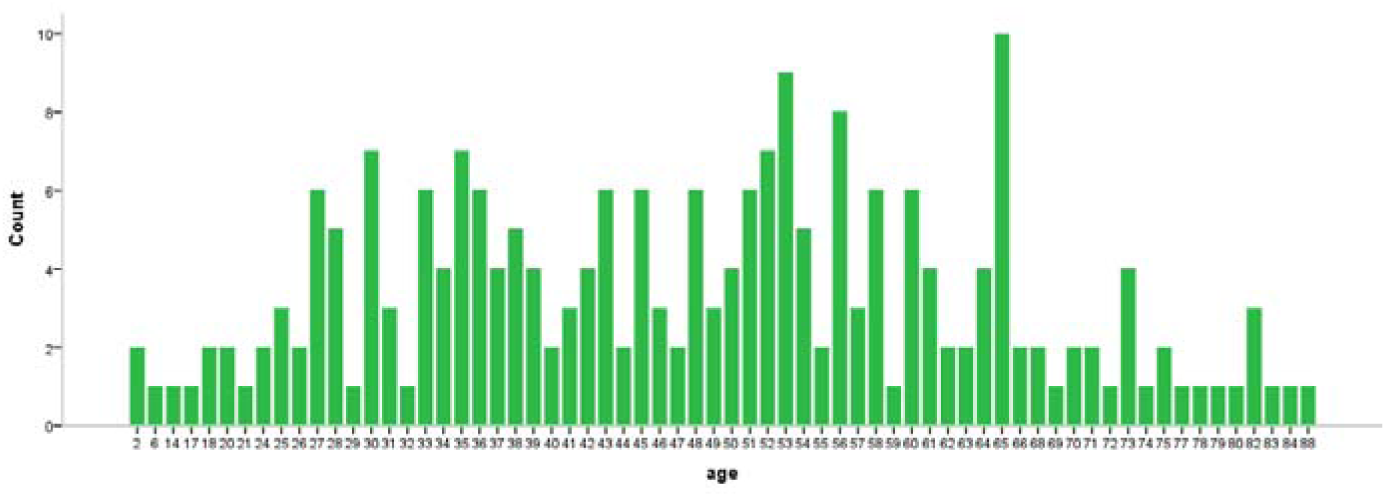
Age distribution of patients with laboratory-confirmed COVID-19

### The epidemiological characteristic

The COVID-19 patients were divided into three groups:

1. Wuhan group (88 cases): Patients with a history of exposure to Wuhan. Sixty-one worked or lived in Wuhan, 22 had short-term exposure by staying in Wuhan for about two days, and five patients only visited a Wuhan railway station or airport for about two hours. These people were free to move around until January 23, and many subsequent cases of infection were caused by the Wuhan group.
2. Family group (62 cases): Patients who had not been to Wuhan within 20 days since the illness started and were infected by acquaintances, mainly family members and other relatives, friends, and colleagues. Some of them only had brief contact with infected people, such as talking or eating together.
3. Stranger group (69 cases): Patients who had not been to Wuhan within the 20 days since the illness started and had been infected by strangers (most of them in a public place). The first patient was a bus driver infected by his passengers; he began to cough and have a fever on January 4. One female patient, who had never been to Wuhan, was a sales clerk in a shopping center. Twenty-three patients were infected in hospital; twelve had uremia and were on dialysis in two hospitals. One citizen was infected only because she went to the supermarket a few times and, since she could not buy a new mask, had been continually wearing the same mask for several days without disinfecting it. Eight medical staff in the non-infected ward were infected by confirmed patients. These staff worked in the gastroenterology department, intensive care unit, emergency department, joint surgery department, and dialysis room. Seven of them became ill between January 21 and 25, and one became ill on January 29.

### The effect of control measures

The number of confirmed patients was analyzed according to the date of symptom onset; for asymptomatic patients, the date of admission was used instead (Figure 2). The number peaked on January 21, which was followed by a gradual decline. Most patients had a history of exposure to Wuhan before January 25, and the number of locally infected cases peaked on January 26. The last patient from Wuhan began to have symptoms of coughing and fever on February 8, after having left Wuhan on January 22. The incubation period was more than 17 days. Most of the patients after February 8, 2020 were family members of confirmed patients. All patients admitted after February 9, 2020 were family members of confirmed patients.

**Figure 2:**
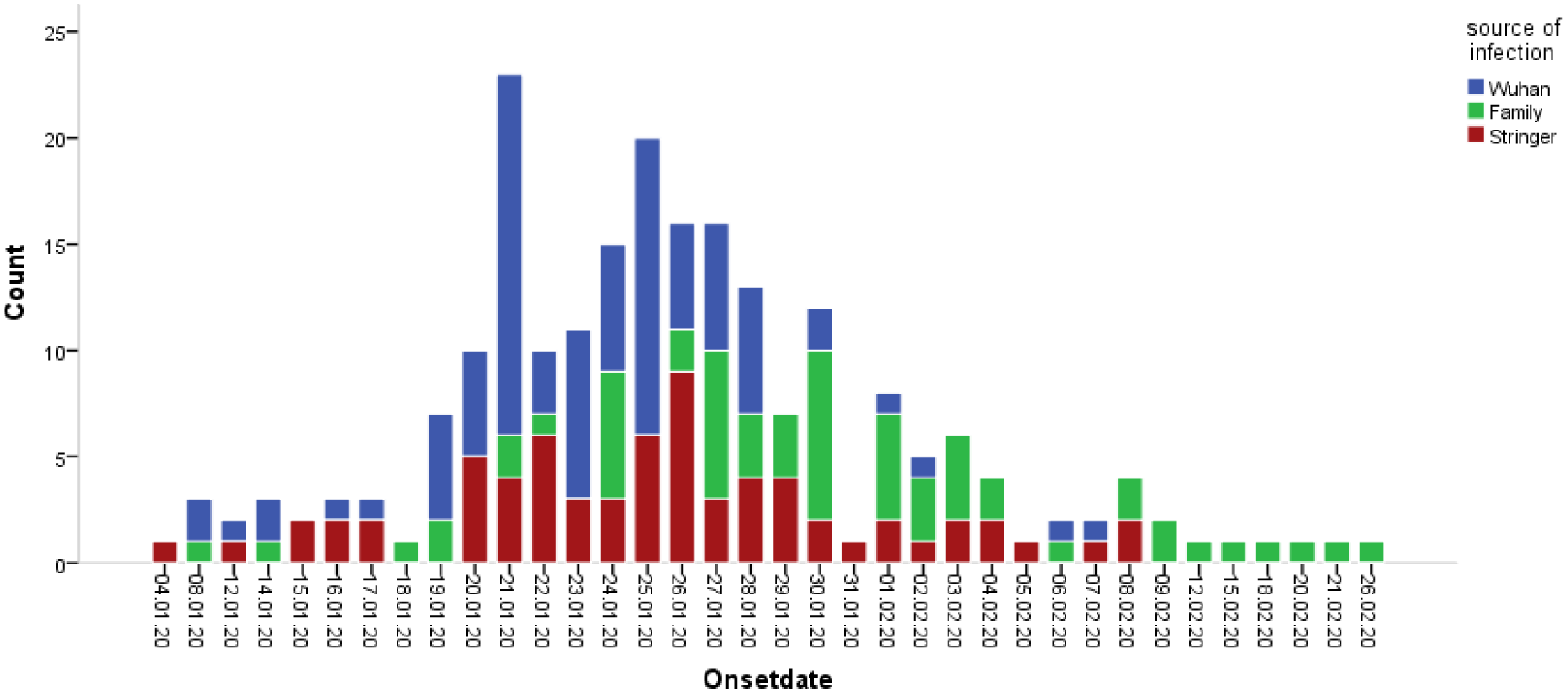
Date of illness onset distribution of patients with COVID-19

## Discussion

SARS-CoV-2 is a newly discovered and named single-strand, positive-sense RNA coronavirus that can cause respiratory disease.^14^ It is related to the other two highly pathogenic viruses, SARS-CoV and MERS-CoV, that have emerged as major global health threats since 2002. SARS-CoV caused 8,422 infections and 800 deaths and spread to 37 countries,^15,16^ and MERS-CoV infected 2,494 individuals and caused 858 deaths worldwide.^17,18^ The number of confirmed COVID-19 patients rose to 70,000 in China by February 20, 2020, with over 44,000 of these in Wuhan. These numbers indicate that SARS-CoV-2 is more contagious than SARS,^19^ and without protection, people can be infected easily when eating together, shopping, meeting, talking, and staying in crowded places.

Most confirmed patients were between the ages of 27 and 68, and there was no difference between genders. These findings are similar to the total cases in China,^20^ which suggests that all humans are sensitive to COVID-19; however, older males and those with comorbid conditions are more likely to have more severe symptoms and potentially die. This is consistent with a previous descriptive study of patients in Jingyintan Hospital, which was responsible for the treatment of serious cases in Wuhan.^10^

No antiviral treatment for COVID-19 has been proven to be effective so far.^21^ The best way to fight the virus is to block its spread. In order to cope with the outbreak of COVID-19, the government of Jingmen has implemented several preventative measures since January 23 (Figure 3). Local residents were advised to stay at home in order to contain the virus spread and were ordered to wear masks in public areas. All markets except supermarkets were closed and all public transportation was suspended. Subsequently, most roads into the city of Jingmen were blocked.

**Figure 3:**
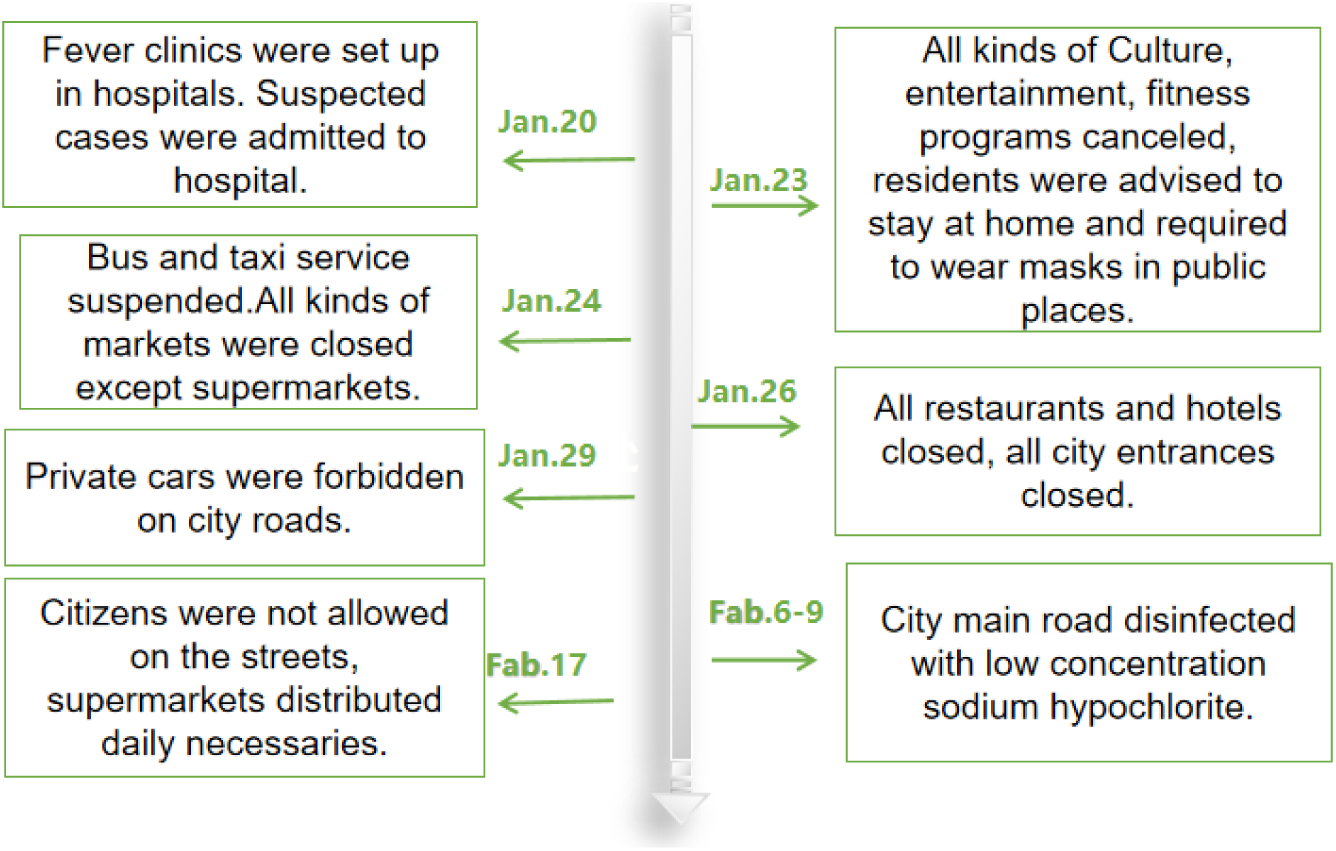
Major prevention and control measures implemented by the goverment of Jingmen (http://www.jingmen.gov.cn)

The benefits from these measures are seen in that the number of locally infected cases peaked on January 26, followed by a gradual decline. Most patients were infected in Wuhan before January 25, and so they could not be controlled.Local infections from strangers were the most dangerous and the hardest to control, since no one knew they were infected before onset of symptoms. There were no new cases infected from strangers after 15 days of the implemention of the control measures, indicating that most of them were infected before the control measures were implemented. The last few cases were family members of confirmed patients, which are hard to prevent when one is living in a home with an infected individual.

More than 20 patients were in the non-infectious ward before their diagnosis. These patients were spread across seven departments, and only eight medical staff members were infected. Moreover, most of them were infected before January 20, when many doctors did not wear masks. Thirteen patients were in the respiratory department before diagnosis; none of the respiratory medical staff had been infected and they had been wearing surgical masks all the time, suggesting that masks can prevent most transmission of COVID-19.

Supermarkets started to limit the number of customers on February 9 and closed on February 17. However, the epidemic had been under control before February 9 (Figure 3), while there were few people infected as everyone wore masks, even though many of them wore only one mask repeatedly without disinfection. This indicated that as long as masks were worn, social gatherings would not cause an outbreak of COVID-19.

Since the time of infection of most patients could not be determined, we did not analyze the incubation period. It was reported that the incubation period was 4–7 days.^22^ Based on the SARS experience, the maximum incubation period is currently considered to be no more than 14 days, but one case in our study was significantly longer at 17 days. A recent study reported the longest incubation period to be 24 days.^11^ The maximum incubation period is unclear.

The asymptomatic case is a challenge to disease control, and 1.2% of cases are asymptomatic in China.^20^ Four asymptomatic cases were diagnosed during screening because they were close contacts and had been hospitalized for more than half a month without symptoms. The pneumonia lesions of these four cases were unilateral and limited to a small area (Figure 4). If they were infected by strangers, they will not be found until some people around them being infected and diagnosed.

There were three limitations in this study. First, only 219 patients diagnosed with COVID-19 were included. Second, the unconfirmed suspected cases in the early stages were excluded in the analyses. Some patients were discharged as suspected patients after one negative nucleic acid test. Many patients had a second or third check before being diagnosed. Third, most of the data were from patients’ medical records, with the potential of a few of them not being accurate enough.

In conclusion, all people are susceptible to COVID-19, and older males and those with comorbid conditions are more likely to become severe cases. Even though COVID-19 is highly contagious, control measures have proven to be very effective, particularly wearing masks, which could prevent most infections.

Declaration of interests

We declare no competing interests

## Data Availability

All data are from the first hospital of Jingmen,if more information are needed, contact the author.

